# Cash transfers may increase the no-show rate for surgical patients in low-resource settings: A randomized trial

**DOI:** 10.1101/2021.03.20.21254039

**Authors:** Mark G. Shrime, Elizabeth A. Harter, Becky Handforth, Christine L. Phillips, Hendrika W.C. Bos, Mirjam Hamer, Dennis Alcorn, Tirzah Bennette, Etienne Faya Millimouno, Jacqueline Nieba, Barry Alpha Oumar, Koko Madeline Zogbe

## Abstract

**Background:** Over two-thirds of the world’s population cannot access surgery when needed. Interventions to address this gap have primarily focused on surgical training and ministry-level surgical planning. However, patients more commonly cite cost— rather than governance or surgeon availability—as their primary access barrier. We undertook a randomized, controlled trial (RCT) to evaluate the effect on compliance with scheduled surgical appointments of addressing this barrier through a cash transfer.

**Methods:** 453 patients who were deemed surgical candidates by a nursing screening team in Guinea, West Africa, were randomized into three study arms: control, conditional cash transfer, and labeled unconditional cash transfer. Arrival to a scheduled surgical appointment was the primary outcome. The study was performed in conjunction with Mercy Ships.

**Results:** The overall no-show rate was five-fold lower in Guinea than previously published estimates, leading to an underpowered study. In a post-hoc analysis, which included non-randomized patients, patients in the control group and the conditional cash transfer group demonstrated no effect from the cash transfer. Patients in the unconditional cash transfer group were significantly less likely to arrive for their scheduled appointment. Subgroup analysis suggested that actual receipt of the unconditional cash transfer, instead of a lapse in the transfer mechanism, was associated with failure to show.

**Conclusion:** We find that cash transfers are feasible for surgical patients in a low-resource setting, but that unconditional transfers may have negative effects on compliance. Although demand-side barriers are large for surgical patients in low-resource settings, interventions to address them must be designed with care.

## INTRODUCTION

Surgical access is critically important for strong health systems.^1^ Although 30% of the world’s disease burden is surgical,^2^ five billion people are unable to access safe, affordable, and timely surgical and anesthesia care.^3^ An estimated 143 million necessary surgical procedures are not done every year.^4^

Barriers to accessing surgical care are especially high among the poor worldwide^5^ and in low- and middle-income countries (LMICs),^3^ where they can put cancer patients at a particularly high risk of poor outcomes.^6^ Many proposals exist to address these barriers. However, most focus on the supply side of the surgical ecosystem: increasing the number of surgeons or non-specialist surgical providers,^7^ the number of surgeries performed by surgeons,^8^ and the quality of these operations.^9^ Other proposals focus on governance of the surgical system, with effort directed toward the development of national surgical, anesthetic, and obstetric plans.^10,11^

These supply-side and governance interventions fail to account for a patient’s ability to get to a surgical appointment. In fact, patients most often cite demand-side barriers, such as cost, rather than supply-side barriers as the primary reason they do not seek surgical care.^12^

Surgical costs can be divided into direct medical costs (such as those for medications, laboratory studies, or the surgery itself), and direct non-medical costs (such as the cost of transportation to the hospital or accommodation). Only 40% of impoverishment from surgery is attributable to direct medical costs; 60% comes from the direct non-medical costs of care,^13^ of which transportation is the largest.

Addressing non-medical costs may increase healthcare utilization.^14^ A 2017 retrospective study of surgical patients in West Africa found that they were almost twice as likely to show up for their scheduled surgery if their transportation costs were paid for.^15^ Vouchers for transportation have also been successful at increasing facility delivery for mothers in Bangladesh, although the voucher system itself proved difficult to administer.^16^

Cash transfers, in which participants are given small amounts of cash in exchange for salutary behavior, are simpler to administer than vouchers and have shown success in health, nutrition, and education.^17^ Cash transfers have not yet been studied in surgery.

Building on a prior modeling study of cash transfers for surgical patients in Guinea,^18^ this paper undertook a randomized, controlled trial (RCT) of a cash transfer for surgical patients in the country. We hypothesized that cash transfers would improve patient compliance and that, specifically, cash transfers given *before* patients faced the barrier of transportation costs would have a significant positive effect.

## METHODS

Mercy Ships is a surgical non-governmental organization that has provided care from the decks of hospital ships since 1978. It focuses primarily in sub-Saharan Africa, where it currently delivers surgery from the m/v *Africa Mercy*. The hospital aboard the *Africa Mercy* has five operating rooms and 75 hospital beds. It has been described in detail elsewhere.^12,15,19^ All surgeries performed on the *Africa Mercy* are elective, and fall into the following surgical specialties: head and neck / maxillofacial surgery, pediatric orthopedics, general and goiter surgery, women’s health, reconstructive surgery, pediatric specialized general surgery, and ophthalmology. (Because ophthalmologic patients are screened through an entirely separate process, they were not included in this RCT.)

Located on the west coast of Africa, Guinea is a country of 95,000 square miles. The country contains 8.3 physicians and 12.4 nurses per 100,000 people in the population.^20^ Conakry, the capital, is home to 1.7 million of the country’s 12.4 million inhabitants.^20^ For ten months beginning in August 2018, the *Africa Mercy* was docked in Conakry.

### Surgical screening

Before they came aboard the *Africa Mercy*, patients were screened for surgical conditions amenable to treatment by the ship hospital. Initial screenings took place at large patient selection events held in five cities throughout Guinea: Boké, Labé, Mamou, Kankan, and N’Zérékoré (**Figure 1**). These cities are between 5 (Boké) and 48 hours (N’Zérékoré) to Conkary by road.

**Figure 1:**
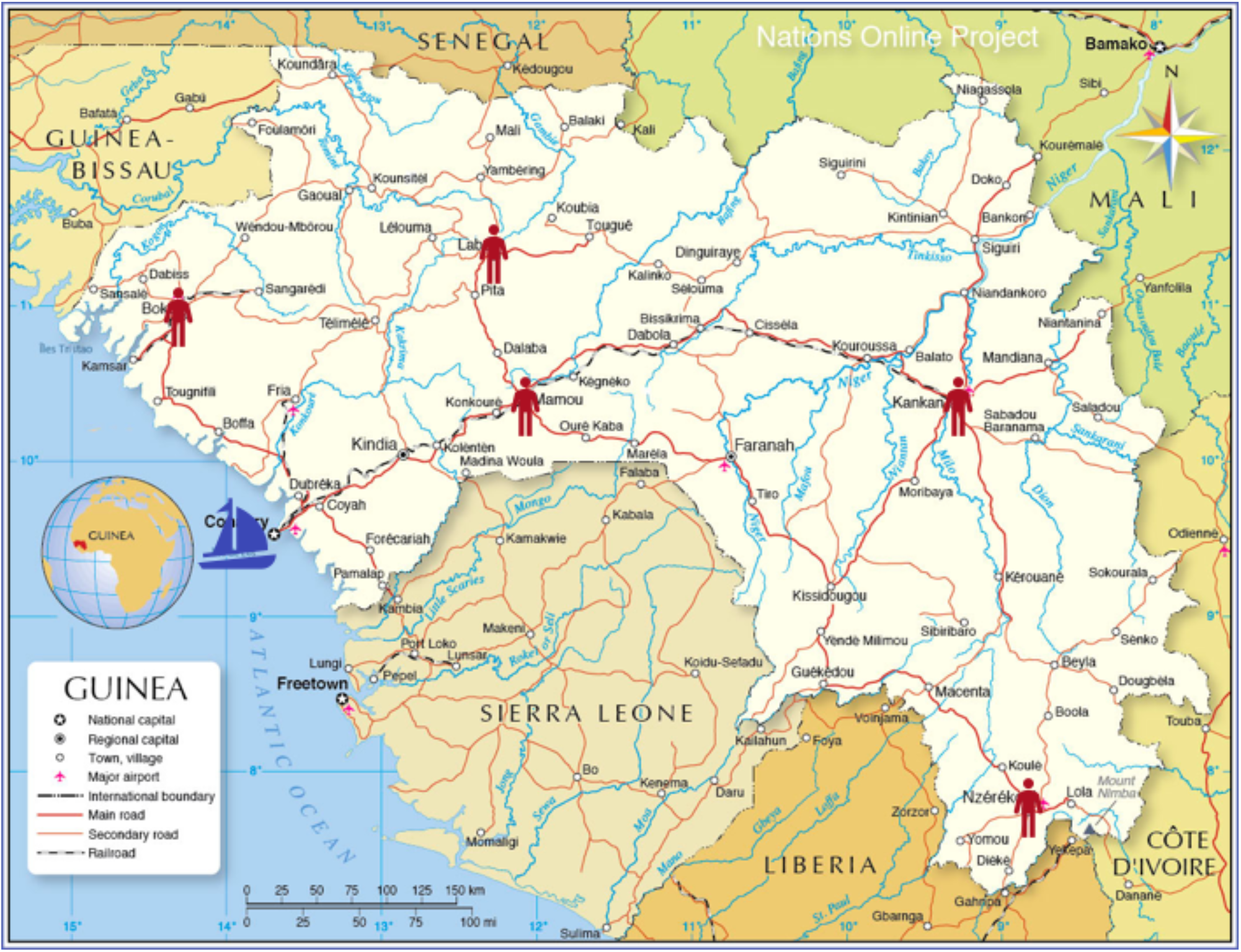
Patient screening sites. Patients were recruited from Boké, Mamou, Labé, Kankan, and N’Zérékoré. They received their surgery on the hospital ship, docked off the coast of Conakry. Map courtesy of the Nations Online Project.

Patients were first seen by a team of up to six volunteer nurses and a dozen Guinean employees. An initial screening selected out those patients with non-surgical complaints (for example, hypertension or diabetes) as their primary presentation, as well as patients with surgical complaints that fell outside the organization’s scope of practice (for example, heart surgery).

Potential surgical patients were then scheduled for diagnostic testing, imaging, and a final surgeon screening, all of which took place on the *Africa Mercy* in Conakry. Once cleared at this stage, patients were scheduled for their operation, usually within two weeks of the final surgeon screening.

For patients selected to travel from field screenings to the second, on-ship screening, Mercy Ships provided transportation from the screening city to Conakry. Patients without a place to stay in or near Conakry were housed, at no cost, in a center established by Mercy Ships. All surgeries, laboratory tests, imaging, medications, and post-operative care were provided by Mercy Ships at no cost to the patient.

Patients scheduled for the on-ship screening were responsible for transportation costs between their homes and the screening city. It is this cost which the RCT was designed to address.

### Study design

The RCT was designed with three arms: a control arm, a conditional cash transfer (CCT) arm, and a labeled, unconditional cash transfer (LUCT) arm. Patients were recruited once they had been given an appointment for their on-ship screening. Allocation proceeded via simple randomization. Slips each containing a random study identification number and group allocation were concealed in an opaque envelope.

Patients drew a number from the envelope in view of the researchers, local staff, and their families. The first author, who performed the data analysis, was blinded to the allocation and randomization. All consents were performed in English with translation into French and the local languages by the Guinean members of the team. Pictograms were used to aid in consent.

CCT patients received the cash transfer conditional on their arrival to the ship. Patients in the LUCT arm received the transfer as a mobile banking deposit 2 – 4 days prior to the day they were scheduled to leave their homes to come to the screening city for transportation to Conakry. The transfer was “labeled” with a concurrent text message informing them that the money was intended to cover their transportation costs. Patients in the control arm received a bag of food and staples on the day of enrollment. No further assistance was given toward their transportation costs.

Given our previous study,^18^ which suggested that an optimal cash transfer is one that offsets all patient costs, and given what Mercy Ships already provided for free, the only cost to be offset by the cash transfer was that for transportation from the patient’s home to their screening city. The average cost of travel from a rural to an urban area was used, estimated to be the no more than or 80,000 Guinean Francs (GNF, approximately $ 9 USD), the price of a one-way ticket from Mamou to Conakry. Patients in the intervention arms received a cash transfer of this amount; the bundle of food and staples provided to patients in the control arm was also worth 80,000 GNF.

The LUCT arm used Orange Money, a widespread mobile banking service throughout Guinea. Patients who did not have a mobile phone were provided one free of charge. Patients who had a mobile phone but did not have an Orange SIM card were provided one free of charge. All patients were given instruction on how the mobile banking transfer would occur.

The study structure is summarized in **Figure 2**, and a CONSORT diagram is included in **Figure 3**.

**Figure 2.**
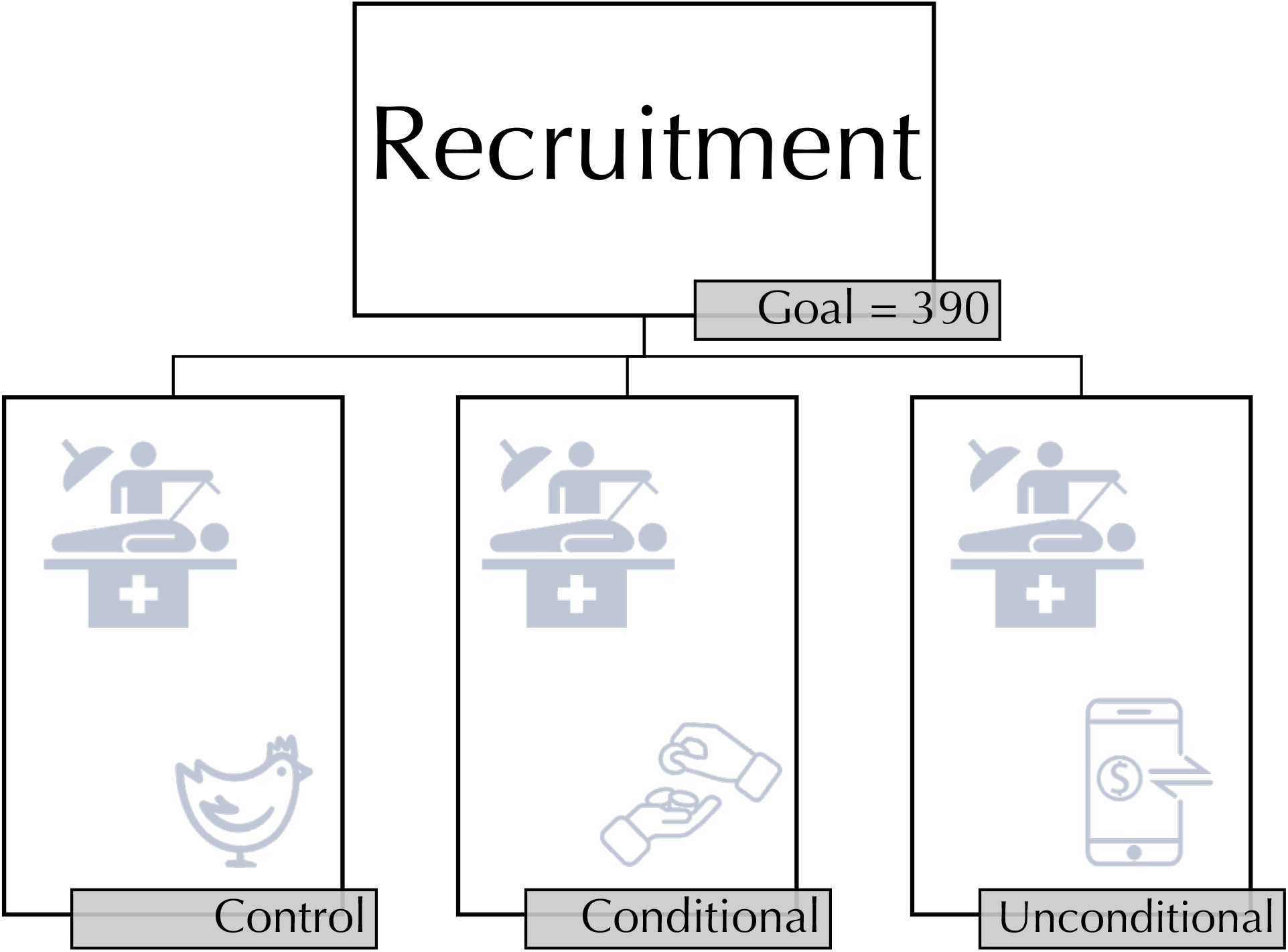
RCT structure. Patients in the control group received a bag of food and staples worth 80,000 GNF (approximately $ 9 USD) on the day of enrolment in the study. Patients in the conditional group received 80,000 GNF in cash at discharge from surgery. Patients in the labeled, unconditional group received 80,000 GNF between two and four days before they were scheduled to leave their homes to arrive for surgery.

**Figure 3.**
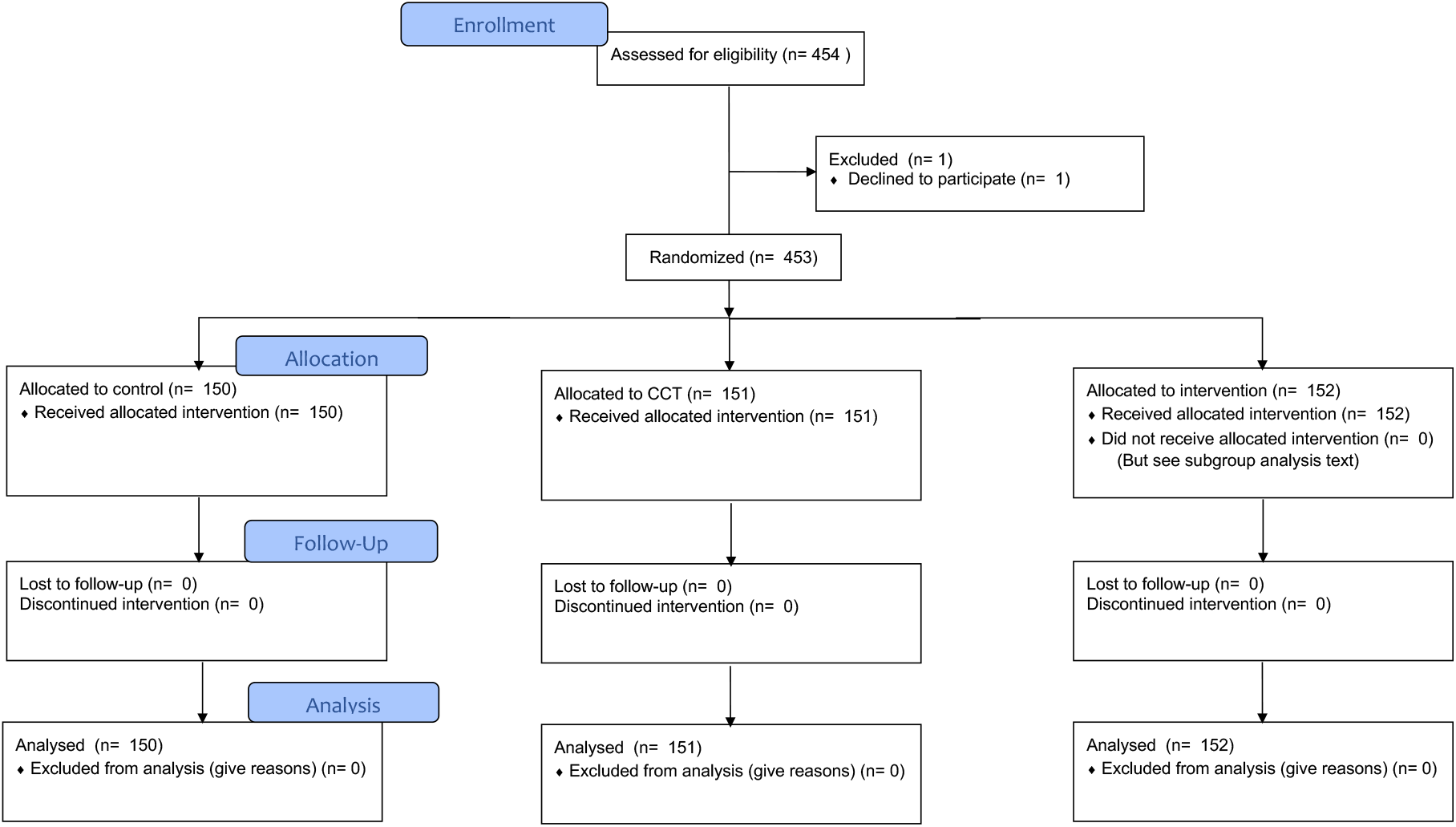
CONSORT diagram

### Sample size and outcomes

Given previously published retrospective data suggesting a decrease in no-show rates from 28% to 15% when transportation is covered,^5^ the sample size for the RCT was calculated at 130 per arm.

The primary outcome of the study was arrival for on-ship screening, not the actual receipt of surgery. Patients were deemed to have complied if they came to this screening—whether or not the surgeon ultimately decided to perform surgical treatment. Patients were deemed non-compliant if they did not arrive for surgeon screening.

Patients in the CCT arm received their transfer irrespective of whether surgery was actually performed. Those who were deemed nonsurgical candidates by the surgeon were given the cash transfer immediately after surgeon screening. Those who had surgery were given the transfer on final discharge.

Analyses were performed in R v3.6.3. Bivariate logistic regression was performed for sufficiently randomized data. For post-hoc analysis with non-randomized data, multivariate logistic regression was performed.

Identifiable data were kept on a secure laptop to which only two members of the study team had access; all analysis was performed blinded. The study was approved by the IRBs of Mercy Ships and of the Massachusetts Eye and Ear Infirmary. The study was funded by the Damon Runyon Cancer Research Foundation (CI-91-17), who had no role in the design, implementation, or reporting of this study. The Guinean Ministry of Health also provided written approval for this study. Study registration: ISRCTN80618786.

## RESULTS

The study successfully enrolled 453 patients, as opposed to the planned 390. Patient demographics are summarized in **Table 1**. No significant difference was noted in observable characteristics among the three study arms. Enrollment for all three groups occurred an average of 67 days prior to their scheduled on-ship screening appointment.

**Table 1:** Demographics of patients included in the randomized trial. CCT = conditional cash transfer. LUCT = labeled unconditional cash transfer.

| **Table 1** | Overall | Control | CCT | LUCT | p |
| --- | --- | --- | --- | --- | --- |
| *Total population* | 453 | 150 | 151 | 152 |  |
| *Specialty, N (%)* |  |  |  |  | 0.659 |
| General | 148 (32.7) | 50 (33.3) | 50 (33.1) | 48 (31.8) |  |
| Maxillofacial / Head and Neck | 176 (38.9) | 59 (39.3) | 56 (37.1) | 61 (40.4) |  |
| Orthopedics | 44 (9.7) | 13 ( 8.7) | 18 (11.9) | 13 ( 8.6) |  |
| Reconstructive surgery | 50 (11.1) | 21 (14.0) | 13 ( 8.6) | 16 (10.6) |  |
| Women's Health | 34 (7.5) | 7 ( 4.7) | 14 ( 9.3) | 13 ( 8.6) |  |
| *Confounders, mean (sd)* |  |  |  |  |  |
| Age | 26.28 (20.17) | 26.19 (20.41) | 24.63 (19.87) | 28.09 (20.24) | 0.402 |
| Male | 0.53 (0.50) | 0.53 (0.50) | 0.53 (0.50) | 0.52 (0.50) | 0.984 |
| Urban wealth quintile | 1.96 (1.09) | 2.04 (1.11) | 1.99 (1.12) | 1.84 (1.03) | 0.287 |
| National wealth quintile | 4.04 (0.98) | 4.09 (0.96) | 3.97 (1.12) | 4.08 (0.86) | 0.553 |
| Travel time (hrs) | 4.51 (10.69) | 3.65 (7.91) | 4.57 (11.88) | 5.32 (11.79) | 0.400 |
| Days between appointments | 67.11 (43.94) | 66.89 (42.89) | 68.81 (44.52) | 65.66 (44.61) | 0.821 |
| *Outcomes, mean (sd)* |  |  |  |  |  |
| Arrived | 0.945 (0.229) | 0.95 (0.212) | 0.96 (0.196) | 0.92 (0.271) | 0.281 |
| Had surgery | 0.845 (0.131) | 0.864 (0.344) | 0.853 (0.355) | 0.819 (0.386) | 0.531 |

The overall no-show rate was significantly lower in Guinea than has been previously reported.^15^ As such, the study was underpowered to detect its primary outcome, as can be seen in **Table 2**.

**Table 2:** Bivariate logistic regression results for the randomized trial. Negative numbers indicate individuals in that group were less likely to return for their scheduled appointment when compared to the control group. CCT = conditional cash transfer. LUCT = labeled unconditional cash transfer.

| *Study patients* | Estimate | Std. Error | *p*-value |
| --- | --- | --- | --- |
| Intercept | 3.0169 | 0.3871 | <0.0001 |
| Control | <Reference> |  |  |
| CCT | 0.168 | 0.5687 | 0.77 |
| LUCT | −0.5602 | 0.4902 | 0.25 |

During the study period, Mercy Ships screened additional patients who were not included in the study. A post-hoc analysis (**Table 3**) compared these patients to those within the study. Because randomization did not occur with these patients, we controlled for all observable confounders for which we had data, as seen in **Table 3**.

**Table 3:** Multivariate logistic regression results, comparing patients within the trial with others screened for surgery by Mercy Ships during the period of the trial. Negative numbers indicate individuals in that group were less likely to return for their scheduled appointment when compared to these out-of-study patients. CCT = conditional cash transfer. LUCT = labeled unconditional cash transfer.

| *Multivariate, all patients* | Estimate | Std. Error | p-value |
| --- | --- | --- | --- |
| (Intercept) | 7.74 | 2.48 | <0.0001 |
| Control | 15.15 | 6403.20 | 1.00 |
| CCT | 15.17 | 6266.95 | 1.00 |
| LUCT | -3.38 | 1.73 | 0.05 |
| Travel Distance | 0.06 | 0.09 | 0.49 |
| Urban wealth quintile | -0.42 | 0.62 | 0.50 |
| Head and neck surgery | -0.23 | 1.45 | 0.88 |
| Orthopedics | 16.53 | 6239.80 | 1.00 |
| Plastics | 16.98 | 5650.42 | 1.00 |
| Women's Health | 17.15 | 8738.50 | 1.00 |

No significant difference in compliance was noted between non-study patients and patients in the control or CCT arms of the study. However, patients in the LUCT arm were significantly less likely to arrive for their surgery (*p* = 0.05), compared with non-study patients.

### Subgroup analysis

For 13.2% of patients in the LUCT group, the transfer could not be completed due to a lack of cell service, a lack of a functioning phone number, or lack of an Orange Money account, despite what the patient provided on enrolment. A subgroup analysis was performed, examining whether the negative association seen in **Table 3** was due to patients who did not receive the cash transfer. The results of this analysis can be seen in **Table 4**: Receipt of the unconditional cash transfer was the only significant predictor of this disincentive (*p* = 0.04).

**Table 4:** Multivariate logistic regression results, comparing patients within the trial with others screened for surgery by Mercy Ships during the period of the trial, and exploring the effect of successful vs. unsuccessful mobile banking transfer. Negative numbers indicate individuals in that group were less likely to return for their scheduled appointment when compared to these out-of-study patients. CCT = conditional cash transfer. LUCT = labeled unconditional cash transfer.

| *Multivariate, all patients* | Estimate | Std. Error | p-value |
| --- | --- | --- | --- |
| (Intercept) | 7.70 | 2.54 | 0.00 |
| Control | 15.15 | 6404.00 | 1.00 |
| CCT | 15.17 | 6269.00 | 1.00 |
| Successful LUCT | −3.49 | 1.72 | 0.04 |
| Unsuccessful LUCT | 15.11 | 18910.00 | 1.00 |
| Travel Distance | 0.07 | 0.09 | 0.48 |
| Urban wealth quintile | −0.41 | 0.64 | 0.52 |
| Maxillofacial surgery | −0.25 | 1.45 | 0.86 |
| Orthopedics | 16.48 | 6218.00 | 1.00 |
| Reconstructive surgery | 16.73 | 5722.00 | 1.00 |
| Women's Health | 17.17 | 8673.00 | 1.00 |

## DISCUSSION

We performed an RCT of a cash transfer to incentivize compliance among patients with surgical disease in Guinea. We found no significant improvement in compliance with patients who received a conditional cash transfer, although the study may have been underpowered to detect this. We did, however, find a significant *decrease* in surgical utilization among patients who received an unconditional cash transfer.

Cash transfers have become an important lever to incentivize salutary behavior in disparate areas such as education, nutrition, HIV treatment, and maternal and child health. Despite their popularity, their results have been mixed.^21–23^ This is, in part, due to the fact that cash transfers are not a monolithic entity. Their design varies across studies, and few studies evaluate the impacts of design choices. Fundamentally, cash can be transferred in two ways: either as a reimbursement *after* the salutary behavior has taken place (a so-called “conditional” cash transfer), or as an incentive prior to the behavior (an “unconditional” cash transfer, in that the transfer is not conditional on the behavior having occurred).

Unconditional cash transfers are less common in global health but have shown some promise. However, recent studies have questioned their effectiveness. A 2017 RCT of cash transfers for maternity care in Kenya suggested that conditionality drove the effect of a cash transfer and that an unconditional transfer “had fewer measured benefits.”^24^

Because transportation is such a significant barrier to access, we hypothesized that the unconditional cash transfer might be the most effective at improving surgical compliance. A conditional transfer would still require patients to face the transportation cost before receiving any reimbursement. Because the trial was performed in conjunction with Mercy Ships, a non-governmental organization that provides surgery for free, covering a patient’s transportation costs meant, for most patients, covering the entirety of the costs they would pay for surgery. Prior modeling has suggested that such coverage would maximize compliance.^18^

That we found the opposite was surprising and leads to an important future direction for research. Three mechanisms may underpin the negative association we observed. The first is that patients simply did not receive the money. **Table 4** implies that this may not be true: patients whom we know did not receive the money in our study were unaffected. What is unknown is what happened to the money after it made it to the recipient’s phone. We do not know if the account holder withdrew the money, nor do we know whether the provided account number actually belonged to the patient, as opposed to a family member or other trusted individual.

A more optimistic explanation for the disincentive would be that that the transfer was large enough that patients received their surgery at a closer medical facility. The average cost of surgery in Guinea is $ 27 – $ 67,^18^ while average Guinean household expenses are $ 53 a month.^20^ A cash transfer of $ 9 is not an insignificant sum.

Finally, and perhaps most believably, the unconditional transfer may have exposed misaligned incentives. The goal of this project was to improve compliance with elective surgery. An unconditional bolus of money may instead have been used to address needs the patient felt to be more pressing, such as food or school fees. This is an area of active research.

This study has some limitations, the most important of which is its power. Although the study was initially powered based on our 2017 published data from surgical care in West Africa, and although the study recruited more patients than initially intended, the no-show rate for surgical patients in Guinea was far lower than that in the literature. Whereas other papers reported double-digit no-show rates, patients presenting to Mercy Ships in Guinea returned for their surgery over 90% of the time, irrespective of group assignment.

This is likely due to the fact that, between the publication of the 2017 paper and the initiation of the trial, the screening team had tripled in size and significant work had been done on understanding the relationship between continual patient contact and compliance. The team’s rapport with patients and retention practices may have contributed to the low no-show rate. Because this study is limited to a single institution, generalization to other settings must be done with caution.

Finally, the control group was not a true control group. Because the Guinean members of the research team felt strongly that randomization to a group that received “nothing” would be perceived poorly, the control group was given a bag of food and staples. Two factors likely mitigate the effect of this intervention. First, the gift was given on the day of enrollment—temporally distant, by over two months on average, from the day of their scheduled surgical appointment. Second, a bag of food and staples is not easily fungible into cash to pay for transportation.

Despite its limitations, this study is the first randomized controlled trial of a behavioral intervention in surgical patients within a resource-constrained setting. It showed the feasibility of such interventions. Further research is needed to understand the mechanism behind the disincentivization created by the unconditional cash transfer.

## CONCLUSION

Cash transfers are a feasible demand-side intervention in global surgery. We find, however, that an unconditional cash transfer in surgery is associated with reduced compliance with scheduled surgical appointments. This highlights the fact that cash transfers should be used with caution, and that attention should be paid specifically to the effects of their design.

## Data Availability

Data are not publicly available

## Notes

### Competing Interest Statement

The authors have declared no competing interest.

### Clinical Trial

ISRCTN80618786

### Author Declarations

The study was approved by the IRBs of Mercy Ships and of the Massachusetts Eye and Ear Infirmary. The Guinean Ministry of Health also provided written approval for this study

